# Clustering of physical health multimorbidity in 68,392 people with severe mental illness and matched comparators: a lifetime prevalence analysis of United Kingdom primary care data

**DOI:** 10.1101/2021.04.30.21256296

**Authors:** Naomi Launders, Joseph F Hayes, Gabriele Price, David PJ Osborn

**Author notes:** Corresponding author Correspondence: Naomi Launders, Division of Psychiatry, UCL. 6^th^ Floor Maple House, 149 Tottenham Court Road, London W1T 7NF. Contributors All authors were involved in the formulated the research question. NL designed the study, analysed the data, and led the writing of the article. DPJO, JFH and GP reviewed the data analysis and contributed to the writing of the article. NL accepts full responsibility for the work and the conduct of the study, had access to the data, and controlled the decision to publish. DPJO and JFH supervised the work. The corresponding author (NL) attests that all listed authors meet authorship criteria and that no others meeting the criteria have been omitted. Copyright The Corresponding Author has the right to grant on behalf of all authors and does grant on behalf of all authors, a worldwide licence to the Publishers and its licensees in perpetuity, in all forms, formats and media (whether known now or created in the future), to i) publish, reproduce, distribute, display and store the Contribution, ii) translate the Contribution into other languages, create adaptations, reprints, include within collections and create summaries, extracts and/or, abstracts of the Contribution, iii) create any other derivative work(s) based on the Contribution, iv) to exploit all subsidiary rights in the Contribution, v) the inclusion of electronic links from the Contribution to third party material where-ever it may be located; and, vi) licence any third party to do any or all of the above. Competing interests All authors have completed the ICMJE uniform disclosure form at www.icmje.org/coi_disclosure.pdf and declare: no support from any commercial organisation for the submitted work; no financial relationships with any organisations that might have an interest in the submitted work in the previous three years; no other relationships or activities that could appear to have influenced the submitted work. Data sharing No additional data are available. Transparency statement The lead author (NL) affirms that the manuscript is an honest, accurate, and transparent account of the study being reported; that no important aspects of the study have been omitted; and that any discrepancies from the study as originally planned have been explained.

## Abstract

**Objective:** To investigate the clustering of physical health multimorbidity in people with severe mental illness (SMI) compared to matched comparators.

**Design:** A cohort-nested analysis of lifetime diagnoses of physical health conditions.

**Setting:** Over 1,800 UK general practices (GP) contributing to Clinical Practice Research DataLink (CPRD) Gold or Aurum databases.

**Participants:** 68,392 adult patients with a diagnosis of SMI between 2000 and 2018, with at least one year of follow up data, matched 1:4 to patients without an SMI diagnosis, on age, sex, GP, and year of GP registration.

**Main outcome measures:** Odds ratios for 24 physical health conditions derived using Elixhauser and Charlson comorbidity indices. We controlled for age, sex, region, and ethnicity; and then additionally for smoking status, alcohol and drug misuse and body mass index. We defined multimorbidity clusters using Multiple Correspondence Analysis and K-Means cluster analysis and described them based on the observed/expected ratio.

**Results:** Patients with a diagnosis of SMI had an increased odds of 19 of 24 physical health conditions and had a higher prevalence of multimorbidity at a younger age compared to comparators (aOR: 2.47; 95%CI: 2.25 to 2.72 in patients aged 20-29). Smoking, obesity, alcohol, and drug misuse were more prevalent in the SMI group and adjusting for these reduced the odds ratio of all comorbid conditions. In patients with multimorbidity (SMI cohort: n=22,843, comparators: n=68,856), we identified six multimorbidity clusters in the SMI cohort, and five in the comparator cohort. Five profiles were common to both. The “hypertension and varied multimorbidity” cluster was most common: 49.8% in the SMI cohort, and 56.7% in comparators. 41.5% of the SMI cohort were in a “respiratory and neurological disease” cluster, compared to 28.7% of comparators.

**Conclusions:** Physical health multimorbidity clusters similarly in people with and without SMI, though patients with SMI develop multimorbidity earlier and a greater proportion fall into a “respiratory and neurological disease” cluster. There is a need for interventions aimed at younger-age multimorbidity in those with SMI.

**Summary box:** *What we already know:*

- People with severe mental illness have higher rates of a range of physical health conditions, including cardiovascular disease, diabetes, and chronic obstructive pulmonary disease (COPD), and a higher mortality rate
- Despite growing attention to disease clustering and profiles of multimorbidity in the general population, there is a lack of evidence regarding multimorbidity clustering in people with SMI.

*What this study adds:*

- Profiles of multimorbidity in people with SMI are similar to the general population, but multimorbidity occurs earlier in those with SMI, with a higher proportion of multimorbid patients defined by clusters of respiratory and neurological disease; services and research should focus on early multimorbidity to decrease the mortality gap, as should commissioners and policy makers.
- People with schizophrenia appear to have lower prevalence of recorded disease for cancer, hypertension, cardiac arrhythmias, valvular disease, and rheumatoid and collagen disease than people without SMI despite high levels of risk factors for these conditions; which requires further investigation to determine whether this is a true effect, or reflects under-diagnosis or inequity in access to healthcare.

## Introduction

People with severe mental illness (SMI) are known to be at increased risk of a range of physical health conditions (1-4), at a younger age(2), and suffer worse outcomes related to these condition (5). The challenges of the increased complexity of managing multiple physical health conditions (6-9) may disproportionally affect those with SMI, further increasing inequality in health outcomes (10, 11) and increasing both secondary mental health and acute service use (12, 13). Comorbidity has been well studied in people with SMI(4). However, while studies have found that people with SMI have a higher number of physical health conditions than the general population (14-19), there is a lack of evidence regarding the clustering of diseases within individuals, or how profiles of physical health multimorbidity in this population compare to those without SMI.

The concept of multimorbidity is of international concern(20), and represents a shift from a single disease-centric approach to a more patient-centred view. The Academy of Medical Science has proposed a definition of multimorbidity which includes long term physical health conditions, infectious diseases of long duration and mental health conditions(21), while the National Institute for Health Care and Excellence (NICE) in England also includes risk factors for disease such as substance misuse(22). There is currently not a common approach to the number or conditions studied, nor the methods used to describe multimorbidity(21, 23-27). However, it is recognised that moving beyond disease pairs or counts of disease is required(26), and that studying the way in which diseases and risk factors cluster within individuals is crucial for improving patient outcomes through better diagnosis, treatment, and healthcare service provision (28, 29). More recently, it has been highlighted that although it is crucial to focus on diagnosing and treating multimorbidity, prevention also requires attention (30).

Mental health diagnoses have been recognised as an important component of multimorbidity in the general population (6, 7, 9, 24, 31-33), but clustering of physical health conditions in patients with SMI is poorly understood. Given the increased disease burden, poorer health outcomes, and higher mortality rate in people with SMI, it is important to characterise the disease profiles occurring in this population. We aimed to investigate the prevalence and clustering of chronic physical health conditions in people with SMI in a large national sample, compared to a matched comparator group without SMI, and investigate the impact of health risk factors in this population

## Methods

### Population

We identified a cohort of patients from the Clinical Practice Research Datalink (CPRD) Gold and Aurum (34, 35) databases. At the time of this study, these databases contained deidentified electronic medical records for over 39 million patients in UK primary care practices. Ethical approval for this study was obtained from the Independent Scientific Advisory Committee of CPRD (protocol no. 18_288).

We included patients with a first diagnosis of SMI between 1 January 2000 and 31 December 2018 via medical codes for schizophrenia, bipolar disorder, or other non-affective psychotic illnesses (code lists available on request). We excluded patients under the age of 18 at SMI diagnosis, and those who had less than one year of active follow up between the age of 18 and 100 years and between 1 January 2000 and 31 December 2018. We matched patients with SMI 1:4 by sex, 5-year age band, primary care practice and year of primary care practice registration with a population of patients without a diagnosis of SMI to generate a comparator cohort.

### Outcomes

The primary outcomes were presence of physical health multimorbidity, defined as two or more of the studied conditions; and lifetime prevalence and odds ratios of these 24 ever-diagnosed chronic physical health conditions in people with SMI compared to the people without SMI. Outcomes were then analysed from the first patient record in the database until the earliest of death, date the patient left the GP practice, or 31 December 2018. We generated code lists for physical health conditions from code lists originally developed by Metcalf et al(36) for calculating the Charlson(37) and Elixhauser(38) comorbidity indices, with a number of modifications specific to considering physical health in people with SMI. We collapsed different severity levels of the same condition into one variable(39) (e.g. uncomplicated diabetes and diabetes with complications were coded as diabetes) and combined the two comorbidity indices into one list. To focus on physical health conditions, we excluded additional diagnoses of psychoses, depression, and dementia(40). Weight loss was defined as a symptom rather than physical health condition and removed from the final analysis. For chronic pulmonary disease, chronic obstructive pulmonary disease (COPD) and asthma were considered as separate conditions.

The final multimorbidity list consisted of 24 conditions: asthma, COPD, cardiac arrhythmia, congestive heart failure, myocardial infarction, cerebrovascular disease, neurological disorders (including epilepsy, multiple sclerosis, Parkinson’s disease and seizures but excluding cerebrovascular disease and dementia), cancer, diabetes (type 1 or 2), hypothyroidism, liver disease, renal disease, peptic ulcers, rheumatic and collagen disease, paresis or paralysis, HIV/AIDS, hypertension, peripheral vascular disease, pulmonary circulation disorders, valvular disease, deficiency anaemia, blood loss anaemia, coagulopathy, and fluid or electrolyte disorders.

### Health risk factors for physical health conditions

We conceptualised alcohol misuse, drug misuse, smoking, and obesity as health risk factors for the development of physical health conditions. We defined alcohol and drug misuse using the code lists for the Elixhauser comorbidity index(36). We categorised BMI as the heaviest ever recorded of obese (BMI≥30), overweight (BMI 25-29.9), healthy weight (BMI 18.5-24.9) or underweight (BMI<18.5), derived from specific medical code lists for obesity, recorded BMI, and BMI calculated from weight and height recording. We categorised smoking status as non-smoker, ex-smoker, or current smoker using medical code lists, taking the most recent category, and recording any non-smokers with a historical code for smoking as ex-smokers.

### Covariates

We defined age as age at the end of follow up based on year of birth, grouped into 10-year age bands. Sex and ethnicity were as recorded in patient medical records and ethnicity was grouped as “Asian”, “Black”, “Mixed”, “White” or “Other”, in line with UK 2011 Census Ethnic Group categories(41). Where multiple ethnicities existed for an individual, we selected the most frequent, and where frequencies were equal, the most recent. Region was based on primary care practice postcode.

### Missing data

As general practitioners are less likely to record values that are within the normal range (42, 43), we coded patients with missing smoking or BMI data as non-smoker or normal range BMI respectively. We coded ethnicity, as recorded in primary care, as white ethnicity where this variable was missing (42). This approach is in line with previous research using primary care data, which suggests that more than 93% of individuals without ethnicity recorded are from a white ethnic group (44).

### Analysis

We determined the prevalence of individual physical health conditions and pairs of conditions (e.g., hypertension and diabetes), stratified by SMI diagnosis, age, and sex. We used logistic regression to investigate the relative prevalence of each physical health condition, first controlling for age, sex, ethnicity, and region and then for these variables plus health risk factors: smoking status, BMI category, alcohol misuse, and drug misuse.

We performed Multiple Correspondence Analysis (MCA) on the population of patients with multimorbidity to investigate the relationship between physical health conditions and to transform the discrete physical health conditions into continuous variables prior to cluster analysis. We then used the MCA dimensions in k-means cluster analysis to identify clusters of physical health conditions and assign individual patients to clusters. Analysis was performed on the subset of patients with multimorbidity; stratified by presence or absence of an SMI diagnosis. We determined the optimum number of clusters by visual inspection of both the Silhouettes(45) and Calinski-Harabaz results(46). We described clusters using the variables with an observed/expected ratio of more than 1.2 or by variables for which more than 70% of patients with that variable were contained within the cluster(24). We then re-ran the MCA and cluster analysis with health risk factors included.

We performed sensitivity analyses to assess the effect of coding missing ethnicity as missing rather than white.

### Patient and public involvement

Patients were not involved at any point of the design or conduct of this study.

## Results

We identified 70,855 patients with a diagnosis of SMI matched to 283,398 comparators. 1,651 patients were excluded as they did not meet the age criteria, 1579 due to less than one year’s follow up, 172 because SMI diagnosis was not within the study period, 1571 because diagnosis was prior to age 18, 145 because of missing practice details and 7,175 because they could not be matched in 1:4 groups. Of the remaining 68,392 patients with SMI, 14,950 had a diagnosis of schizophrenia, 24,328 a diagnosis of bipolar disorder, and 29,114 a diagnosis of other psychoses. A higher proportion of patients with SMI died between 2000 and 2018 than those without SMI and death occurred at a younger mean age (Table 1). A greater proportion of patients in the comparator group had missing information for ethnicity (45.8% vs. 38.2%; supplementary table 1), smoking (7.0% vs. 2.3%) and BMI (18.8% vs. 9.8%) than in the SMI cohort.

**Table 1:** Characteristics of the SMI cohort and matched comparators, n=341,960.

|  | No SMI | SMI |  |  |  |
| --- | --- | --- | --- | --- | --- |
|  |  | All SMI | Schizophrenia | Bipolar | Other |
| **Number of patients** | 273,568 | 68,392 | 14,950 | 24,328 | 29,114 |
| **Age at end of study (mean (SD))** | 50.62 (19.53) | 50.7 (18.98) | 48.27 (17.25) | 50.57 (17.23) | 52.04 (20.98) |
| **Age at diagnosis (mean (SD))<sup>a</sup>** | 44.64 (18.71) | 44.64 (18.71) | 41.57 (17.00) | 44.24 (16.41) | 46.55 (20.98) |
| **Female (%)** | 134164 (49.0) | 33541 (49.0) | 5314 (35.5) | 14397 (59.2) | 13830 (47.5) |
| **Ethnicity (%)<sup>b</sup>** |  |  |  |  |  |
| Asian | 13794 (5.0) | 3230 (4.7) | 1057 (7.1) | 777 (3.2) | 1396 (4.8) |
| Black | 8899 (3.3) | 3593 (5.3) | 1428 (9.6) | 534 (2.2) | 1631 (5.6) |
| Mixed | 2101 (0.8) | 831 (1.2) | 237 (1.6) | 249 (1.0) | 345 (1.2) |
| Other | 6151 (2.2) | 1579 (2.3) | 354 (2.4) | 537 (2.2) | 688 (2.4) |
| White | 242623 (88.7) | 59159 (86.5) | 11874 (79.4) | 22231 (91.4) | 25054 (86.1) |
| **BMI category (%)<sup>c</sup>** |  |  |  |  |  |
| Underweight | 4244 (1.6) | 790 (1.2) | 197 (1.3) | 168 (0.7) | 425 (1.5) |
| Healthy weight | 126520 (46.2) | 22431 (32.8) | 4838 (32.4) | 6744 (27.7) | 10849 (37.3) |
| Overweight | 74534 (27.2) | 19070 (27.9) | 3905 (26.1) | 7015 (28.8) | 8150 (28.0) |
| Obese | 68270 (25.0) | 26101 (38.2) | 6010 (40.2) | 10401 (42.8) | 9690 (33.3) |
| **Smoking category (%)<sup>d</sup>** |  |  |  |  |  |
| Current smoker | 75159 (27.5) | 31371 (45.9) | 8009 (53.6) | 10281 (42.3) | 13081 (44.9) |
| Ex-smoker | 85303 (31.2) | 20593 (30.1) | 3605 (24.1) | 8329 (34.2) | 8659 (29.7) |
| Non smoker | 113106 (41.3) | 16428 (24.0) | 3336 (22.3) | 5718 (23.5) | 7374 (25.3) |
| **Alcohol misuse (%)** | 8782 (3.2) | 8659 (12.7) | 1751 (11.7) | 3023 (12.4) | 3885 (13.3) |
| **Drug misuse (%)** | 5609 (2.1) | 8547 (12.5) | 2096 (14.0) | 2260 (9.3) | 4191 (14.4) |
| **Physical health conditions** |  |  |  |  |  |
| No physical health conditions | 139537 (51.0) | 27134 (39.7) | 6789 (45.4) | 8968 (36.9) | 11377 (39.1) |
| One physical health condition | 65175 (23.8) | 18415 (26.9) | 3974 (26.6) | 6906 (28.4) | 7535 (25.9) |
| More than one physical health condition | 68856 (25.2) | 22843 (33.4) | 4187 (28.0) | 8454 (34.8) | 10202 (35.0) |
| **Deaths (%)** | 19387 (7.1) | 7514 (11.0) | 1606 (10.7) | 2148 (8.8) | 3760 (12.9) |
| **Age at death (mean, SD)** | 78.94 (14.35) | 71.43 (18.05) | 66.01 (18.08) | 70.09 (15.81) | 74.52 (18.59) |
| **Years of observation (mean, SD)** | 30.72 (16.87) | 30.53 (16.62) | 25.91 (16.13) | 32.03 (16.13) | 31.66 (16.85) |

SD: Standard deviation; a: For comparator population, this is the age of the individual at the time of diagnosis for the corresponding case; b: Missing ethnicity set to white (n=151,320, 44.3%), c: missing BMI set to normal (n=58,033, 17.0%); d: missing smoking set to non-smoker (n=20,765, 6.1%).

### Prevalence of chronic physical health conditions and multimorbidity

There was a higher prevalence of at least one physical health condition (SMI: 60.3%; 95% confidence interval [CI]: 60.0 to 60.7%, comparator: 49.0%; 95%CI: 48.8 to 49.2%) and multimorbidity in the SMI cohort (Table 1). When controlling for age, sex, ethnicity, and region those with SMI were at increased risk of multimorbidity (adjusted odds ratio [aOR]: 1.81 (95%CI: 1.78 to 1.85). Additionally controlling for smoking status, BMI category, alcohol misuse, and drug misuse reduced the odds ratio for multimorbidity in patients with SMI, though it was still elevated (aOR: 1.63; 95%CI: 1.57 to 1.70). In both cohorts multimorbidity was more common in females and in older age groups (Figure 1), though the greatest difference in prevalence of multimorbidity between those with and without SMI was in patients aged 20-29 (aOR: 2.47; 95%CI: 2.25 to 2.72) and the difference got smaller with increasing age. At age 80-89 prevalence of multimorbidity was similar in both cohorts (supplementary table 2).

**Figure 1:**
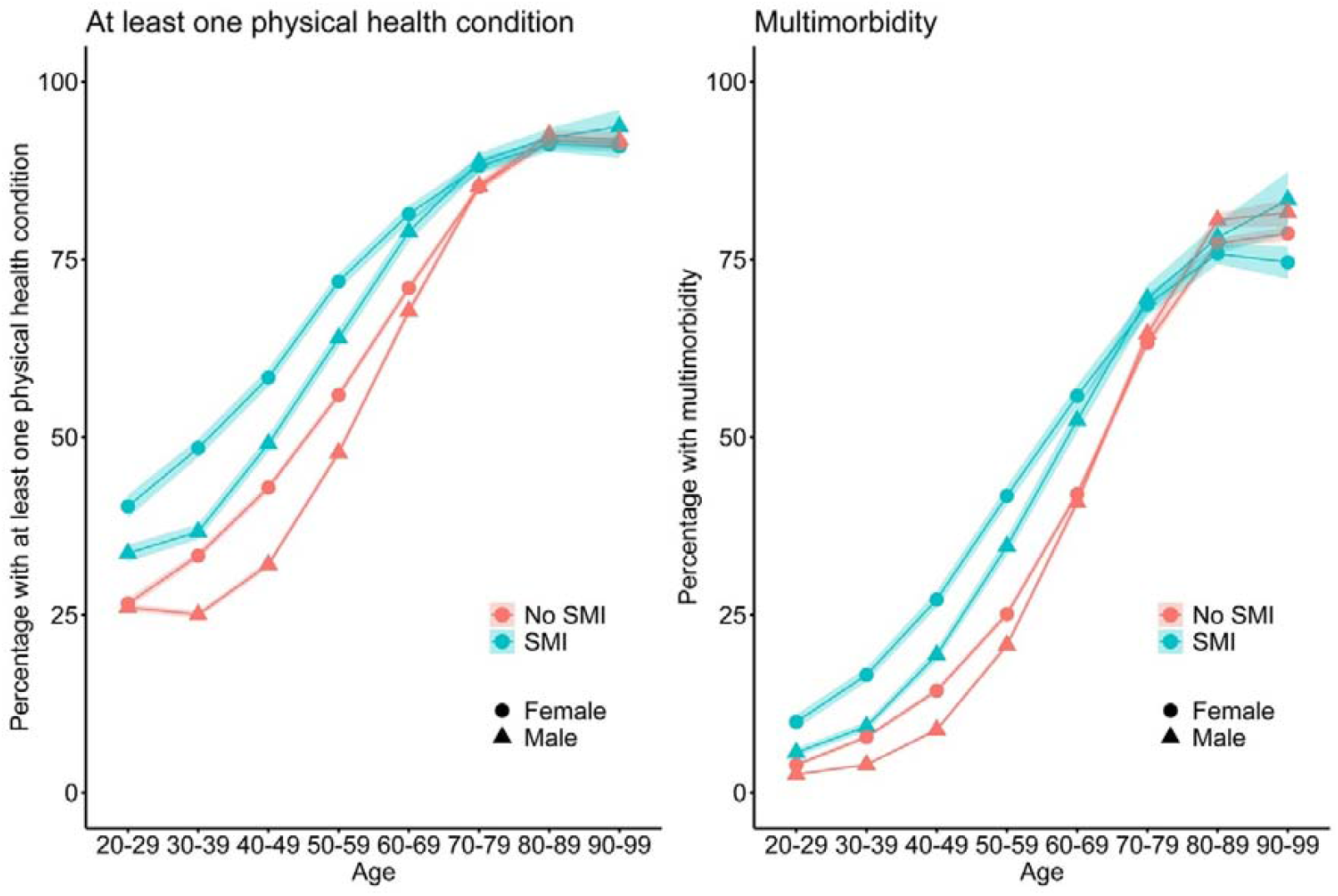
Prevalence of at least one physical health condition and multimorbidity, by SMI status, age group and sex.

The most common physical health conditions were hypertension, asthma, and diabetes in both SMI and non-SMI cohorts, and when stratified by SMI diagnosis (Table 2). The most common multimorbidity pairs were the same in those with and without SMI: hypertension and diabetes (SMI: 7.2%; no SMI: 6.0%) followed by hypertension and renal disease (SMI: 4.6%; no SMI: 4.9%; Figure 2).

**Table 2:** Prevalence of physical health conditions.

|  | Prevalence, n, (%) |  |  |  |  | Partially adjusted, Odds Ratio (95% CI) |  |  |  | Fully adjusted, Odds Ratio (95% CI) |  |  |  |
| --- | --- | --- | --- | --- | --- | --- | --- | --- | --- | --- | --- | --- | --- |
|  | No SMI | SMI | Schiz | Bipolar | Other | SMI | Schiz | Bipolar | Other | SMI | Schiz | Bipolar | Other |
| Asthma | 41596 (15.2) | 12681 (18.5) | 2336 (15.6) | 5109 (21.0) | 5236 (18.0) | 1.28 (1.25-1.31) | 1.07 (1.02-1.12) | 1.45 (1.40-1.49) | 1.24 (1.20-1.28) | 1.06 (1.03-1.08) | 0.87 (0.83-0.91) | 1.17 (1.13-1.21) | 1.06 (1.02-1.09) |
| COPD | 10017 (3.7) | 3739 (5.5) | 771 (5.2) | 1399 (5.8) | 1569 (5.4) | 1.62 (1.55-1.68) | 1.82 (1.68-1.97) | 1.89 (1.78-2.01) | 1.35 (1.28-1.43) | 1.21 (1.16-1.26) | 1.27 (1.17-1.38) | 1.36 (1.28-1.44) | 1.08 (1.02-1.15) |
| Cardiac arrhythmia | 14329 (5.2) | 3632 (5.3) | 541 (3.6) | 1167 (4.8) | 1924 (6.6) | 1.07 (1.02-1.11) | 0.91 (0.83-0.99) | 1.11 (1.04-1.18) | 1.10 (1.04-1.16) | 1.00 (0.96-1.04) | 0.86 (0.78-0.94) | 0.98 (0.92-1.05) | 1.06 (1.01-1.12) |
| Congestive heart failure | 6996 (2.6) | 1871 (2.7) | 288 (1.9) | 580 (2.4) | 1003 (3.4) | 1.15 (1.08-1.21) | 1.06 (0.94-1.20) | 1.27 (1.16-1.39) | 1.11 (1.03-1.19) | 1.07 (1.01-1.13) | 0.99 (0.87-1.12) | 1.11 (1.01-1.21) | 1.08 (1.00-1.16) |
| Myocardial infarction | 6186 (2.3) | 1578 (2.3) | 252 (1.7) | 459 (1.9) | 867 (3.0) | 1.08 (1.02-1.14) | 0.96 (0.84-1.10) | 1.02 (0.93-1.13) | 1.15 (1.07-1.24) | 0.99 (0.94-1.05) | 0.87 (0.76-0.99) | 0.89 (0.80-0.98) | 1.12 (1.03-1.20) |
| Cerebrovascular disease | 11703 (4.3) | 3959 (5.8) | 585 (3.9) | 1335 (5.5) | 2039 (7.0) | 1.51 (1.45-1.57) | 1.28 (1.17-1.40) | 1.71 (1.60-1.82) | 1.47 (1.40-1.55) | 1.40 (1.35-1.46) | 1.18 (1.07-1.29) | 1.53 (1.44-1.63) | 1.41 (1.33-1.48) |
| Neurological disease | 6320 (2.3) | 4159 (6.1) | 879 (5.9) | 1370 (5.6) | 1910 (6.6) | 2.76 (2.65-2.87) | 2.78 (2.59-2.99) | 2.51 (2.37-2.67) | 2.95 (2.80-3.11) | 2.41 (2.31-2.52) | 2.44 (2.26-2.63) | 2.18 (2.05-2.32) | 2.60 (2.46-2.74) |
| Cancer | 23828 (8.7) | 5442 (8.0) | 838 (5.6) | 2060 (8.5) | 2544 (8.7) | 0.92 (0.89-0.96) | 0.80 (0.74-0.86) | 1.11 (1.05-1.16) | 0.85 (0.81-0.89) | 0.89 (0.86-0.92) | 0.77 (0.72-0.83) | 1.03 (0.98-1.08) | 0.83 (0.79-0.87) |
| Diabetes | 29673 (10.8) | 11956 (17.5) | 2923 (19.6) | 4276 (17.6) | 4757 (16.3) | 1.81 (1.77-1.86) | 2.11 (2.02-2.21) | 2.06 (1.98-2.14) | 1.51 (1.45-1.56) | 1.47 (1.43-1.51) | 1.66 (1.58-1.74) | 1.51 (1.45-1.57) | 1.34 (1.29-1.39) |
| Hypothyroidism | 13180 (4.8) | 5438 (8.0) | 789 (5.3) | 2659 (10.9) | 1990 (6.8) | 1.81 (1.74-1.87) | 1.50 (1.39-1.62) | 2.58 (2.46-2.70) | 1.35 (1.28-1.42) | 1.68 (1.62-1.74) | 1.39 (1.28-1.50) | 2.28 (2.17-2.39) | 1.32 (1.25-1.39) |
| Liver disease | 3987 (1.5) | 1923 (2.8) | 445 (3.0) | 572 (2.4) | 906 (3.1) | 1.94 (1.83-2.05) | 1.96 (1.77-2.16) | 1.72 (1.58-1.88) | 2.09 (1.95-2.25) | 1.06 (1.00-1.13) | 1.08 (0.97-1.20) | 0.97 (0.88-1.07) | 1.12 (1.03-1.21) |
| Renal disease | 18225 (6.7) | 5294 (7.7) | 888 (5.9) | 1934 (7.9) | 2472 (8.5) | 1.27 (1.23-1.32) | 1.24 (1.14-1.33) | 1.62 (1.54-1.71) | 1.07 (1.02-1.13) | 1.20 (1.16-1.24) | 1.17 (1.08-1.27) | 1.41 (1.34-1.49) | 1.07 (1.02-1.13) |
| Peptic ulcer | 5702 (2.1) | 1713 (2.5) | 248 (1.7) | 580 (2.4) | 885 (3.0) | 1.24 (1.17-1.31) | 0.90 (0.79-1.02) | 1.32 (1.21-1.45) | 1.33 (1.24-1.43) | 1.05 (0.99-1.11) | 0.74 (0.64-0.84) | 1.09 (1.00-1.20) | 1.16 (1.07-1.24) |
| Rheumatic/collagen diseases | 7452 (2.7) | 1794 (2.6) | 227 (1.5) | 692 (2.8) | 875 (3.0) | 0.98 (0.93-1.03) | 0.71 (0.62-0.81) | 1.14 (1.05-1.23) | 0.97 (0.90-1.04) | 0.93 (0.89-0.99) | 0.67 (0.59-0.77) | 1.04 (0.96-1.13) | 0.95 (0.89-1.03) |
| Paresis/paralysis | 1235 (0.5) | 602 (0.9) | 92 (0.6) | 219 (0.9) | 291 (1.0) | 1.97 (1.78-2.17) | 1.44 (1.16-1.78) | 2.04 (1.76-2.35) | 2.16 (1.90-2.46) | 1.90 (1.72-2.11) | 1.39 (1.12-1.73) | 1.94 (1.67-2.25) | 2.11 (1.85-2.41) |
| HIV | 1755 (0.6) | 745 (1.1) | 115 (0.8) | 297 (1.2) | 333 (1.1) | 1.66 (1.52-1.81) | 1.13 (0.93-1.37) | 1.83 (1.61-2.07) | 1.80 (1.59-2.02) | 1.26 (1.14-1.38) | 0.85 (0.69-1.03) | 1.39 (1.22-1.59) | 1.35 (1.19-1.53) |
| Hypertension | 54693 (20.0) | 13404 (19.6) | 2386 (16.0) | 4835 (19.9) | 6183 (21.2) | 0.97 (0.95-0.999) | 0.87 (0.82-0.91) | 1.15 (1.10-1.19) | 0.89 (0.86-0.93) | 0.79 (0.77-0.82) | 0.70 (0.66-0.74) | 0.84 (0.81-0.88) | 0.80 (0.77-0.83) |
| Peripheral vascular | 5388 | 1439 | 219 | 494 | 726 | 1.13 (1.06- | 0.96 (0.83- | 1.29 (1.17- | 1.09 (1.01- | 0.98 (0.92- | 0.79 (0.68- | 1.06 (0.97- | 0.99 (0.91- |

| disease | (2.0) | (2.1) | (1.5) | (2.0) | (2.5) | 1.20) | 1.10) | 1.42) | 1.18) | 1.04) | 0.91) | 1.17) | 1.07) |
| --- | --- | --- | --- | --- | --- | --- | --- | --- | --- | --- | --- | --- | --- |
| Pulmonary circulation disorders | 2836<br>(1.0) | 1043<br>(1.5) | 171<br>(1.1) | 382<br>(1.6) | 490<br>(1.7) | 1.52 (1.41-1.63) | 1.38 (1.18-1.62) | 1.70 (1.52-1.89) | 1.44 (1.31-1.59) | 1.31 (1.22-1.41) | 1.18 (1.00-1.38) | 1.37 (1.23-1.53) | 1.32 (1.19-1.45) |
| Valvular disease | 4883<br>(1.8) | 1125<br>(1.6) | 153<br>(1.0) | 379<br>(1.6) | 593<br>(2.0) | 0.95 (0.89-1.01) | 0.73 (0.62-0.86) | 1.03 (0.92-1.14) | 0.97 (0.89-1.06) | 0.91 (0.85-0.98) | 0.71 (0.60-0.84) | 0.95 (0.85-1.05) | 0.96 (0.88-1.05) |
| Deficiency anaemia | 14248<br>(5.2) | 4763<br>(7.0) | 885<br>(5.9) | 1690<br>(6.9) | 2188<br>(7.5) | 1.37 (1.32-1.42) | 1.33 (1.24-1.43) | 1.39 (1.31-1.46) | 1.37 (1.31-1.44) | 1.30 (1.25-1.35) | 1.26 (1.18-1.36) | 1.27 (1.21-1.34) | 1.33 (1.27-1.40) |
| Blood loss anaemia | 185<br>(0.1) | 43<br>(0.1) | 11<br>(0.1) | 14<br>(0.1) | 18<br>(0.1) | 0.90 (0.64-1.25) | 1.17 (0.63-2.16) | 0.87 (0.50-1.50) | 0.80 (0.49-1.30) | 0.87 (0.62-1.22) | 1.11 (0.60-2.07) | 0.82 (0.47-1.42) | 0.80 (0.49-1.30) |
| Coagulopathy | 1786<br>(0.7) | 612<br>(0.9) | 100<br>(0.7) | 224<br>(0.9) | 288<br>(1.0) | 1.38 (1.25-1.51) | 1.08 (0.88-1.32) | 1.48 (1.28-1.70) | 1.43 (1.27-1.63) | 1.16 (1.05-1.28) | 0.91 (0.74-1.12) | 1.20 (1.04-1.39) | 1.24 (1.09-1.41) |
| Fluid and electrolyte disorders | 5314<br>(1.9) | 2291<br>(3.3) | 352<br>(2.4) | 859<br>(3.5) | 1080<br>(3.7) | 1.85 (1.76-1.95) | 1.58 (1.41-1.77) | 2.27 (2.11-2.45) | 1.68 (1.57-1.80) | 1.62 (1.54-1.71) | 1.38 (1.24-1.55) | 1.88 (1.74-2.03) | 1.54 (1.44-1.65) |

Partially adjusted: Adjusted for age, sex, ethnicity, and region.
Fully adjusted: Adjusted for age, sex, ethnicity, and region, and additionally for smoking status, BMI category, alcohol, and drug misuse.

**Figure 2:**
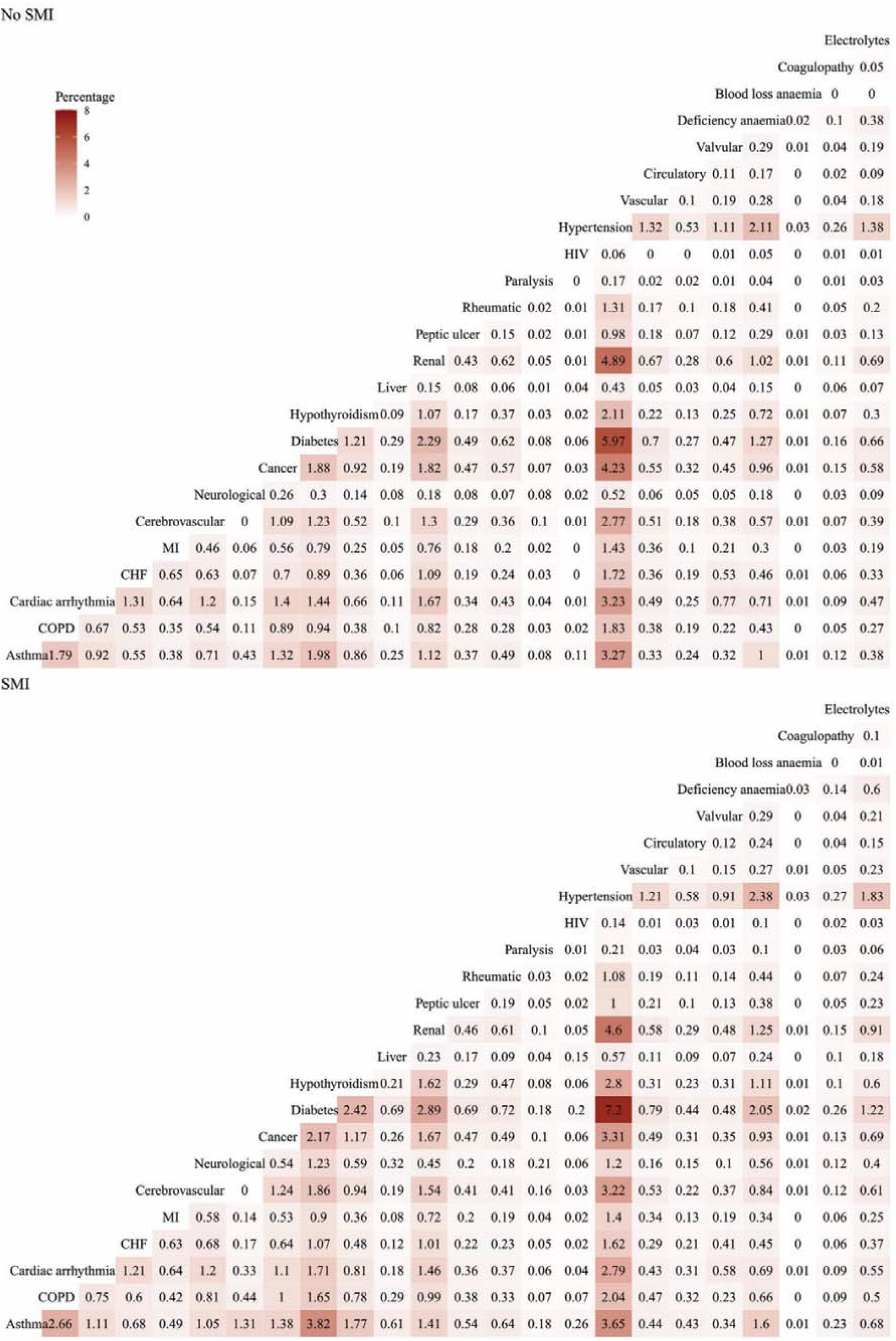
Percentage of patients with each combination of physical health condition pairs in patients with SMI and matched comparators. CHF: congestive heart failure, MI: myocardial infarction; Rheumatic: rheumatic and collagen disease; Paralysis: paralysis/paresis; Vascular: peripheral vascular disease; circulatory: pulmonary circulation disorders; Electrolytes: Fluid and electrolyte disorders

When adjusting for age, sex, ethnicity and region, patients with SMI had greater odds of recorded diagnoses of 19 out of 24 diseases (Figure 3; Table 2). When stratified by SMI diagnosis, patients with schizophrenia had lower odds of recorded cardiac arrythmia, cancer, valvular disease, rheumatoid and collagen disease, and hypertension than the comparator population (Table 2).

**Figure 3:**
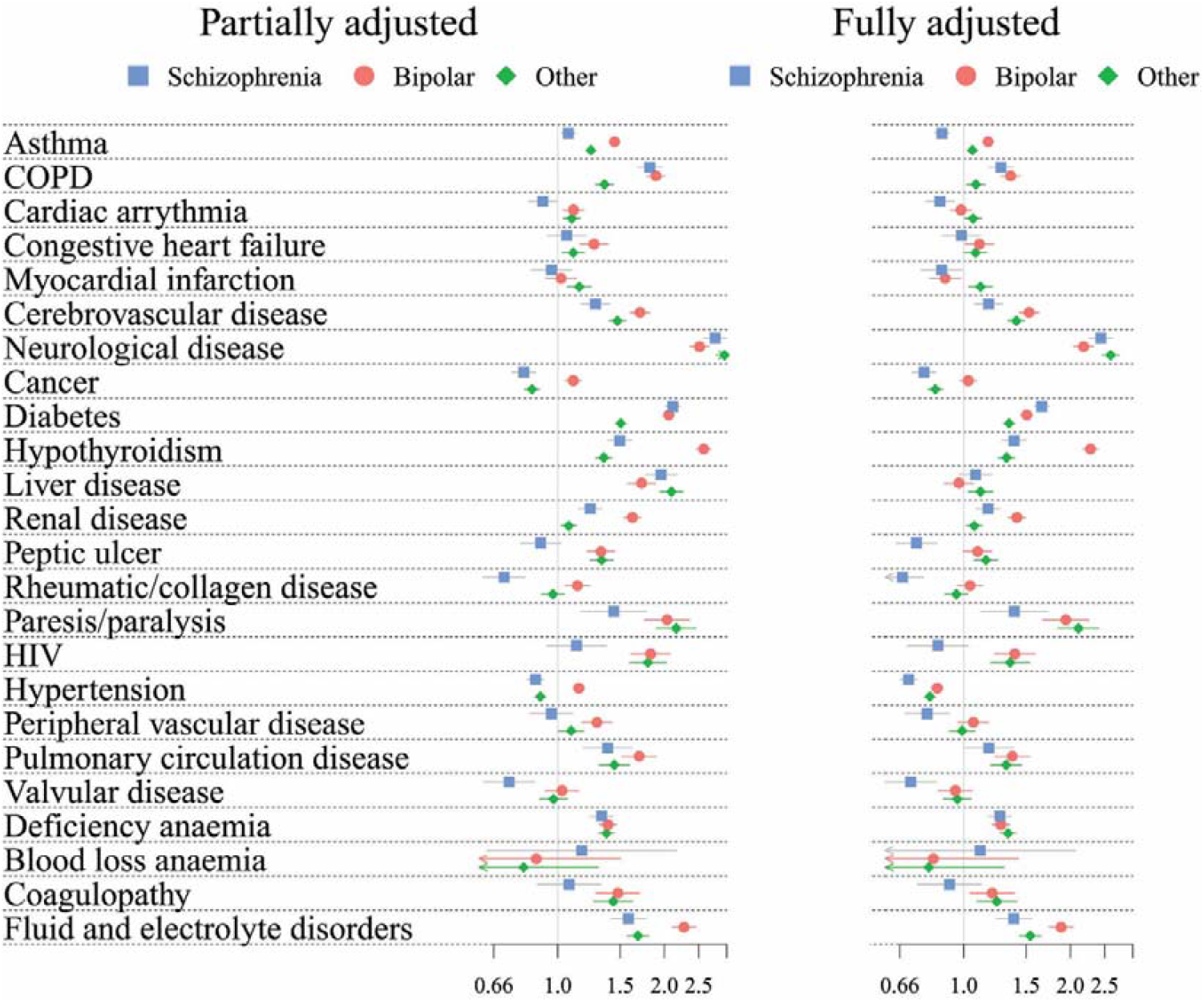
Odds ratios of physical health conditions in those with SMI compared to the comparator population.

Patients with SMI had more health risk factors than the comparator population (Table 1). Obesity was particularly prevalent in those with a diagnosis of bipolar disorder (42.8%), while smoking was most prevalent in those with schizophrenia (53.6%), and alcohol and drug misuse most prevalent in those with a diagnosis of other psychoses (13.3% and 14.4% respectively). After adjustment for these risk factors the odds ratios for all diseases reduced, in particular for liver disease, COPD, HIV, diabetes and hypertension (Figure 3; Table 2).

### Clustering of physical health conditions and multimorbidity profiles

In multiple correspondence analysis of patients with physical health multimorbidity (SMI cohort:, comparators:, 16 dimensions were required to explain 70% of the variance of physical health conditions in the SMI cohort and 15 in the comparator cohort. The first two dimensions in MCA had similar disease profiles, with the first defined by heart disease and the second by respiratory disease for both cohorts (Supplementary figure 1).

We identified six profiles of physical health multimorbidity using K-means cluster analysis of the MCA dimensions. All clusters were present in the cohort of patients with SMI and all except a small cluster of patients with blood loss anaemia (SMI: n=35), were present in the comparator cohort. The largest patient group (49.84% of the SMI population, 56.68.0% of the non-SMI population) consisted of patients with a high prevalence of hypertension, combined with a variety of other health conditions (Table 3, Figure 4); the “hypertension and varied multimorbidity cluster”. Patients with SMI were less likely than the comparator cohort to fall into this and the “heart disease” cluster (SMI: 4.5%; comparator: 10.0%). Both these clusters were characterised by older age and low prevalence of health risk factors.

**Table 3:**
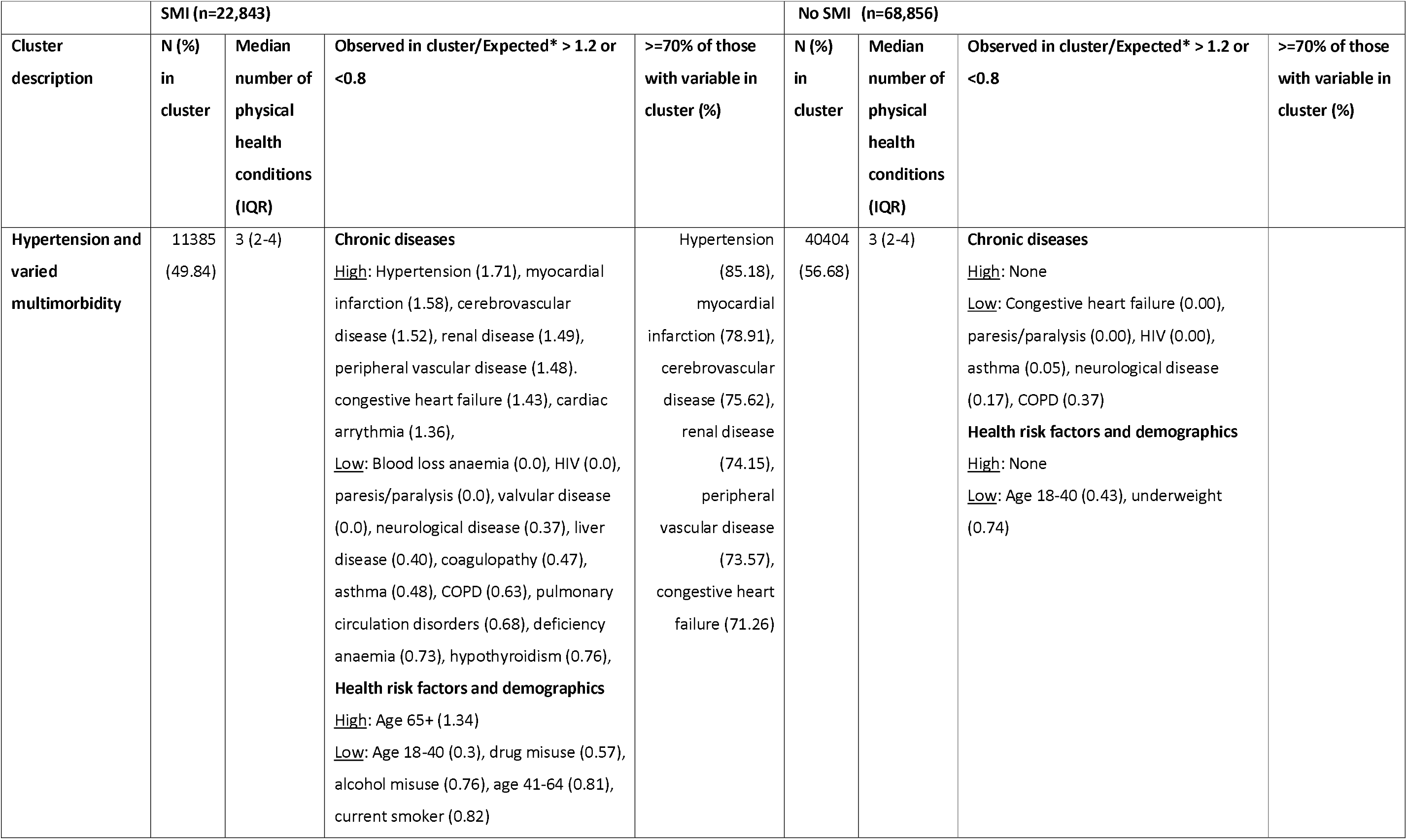
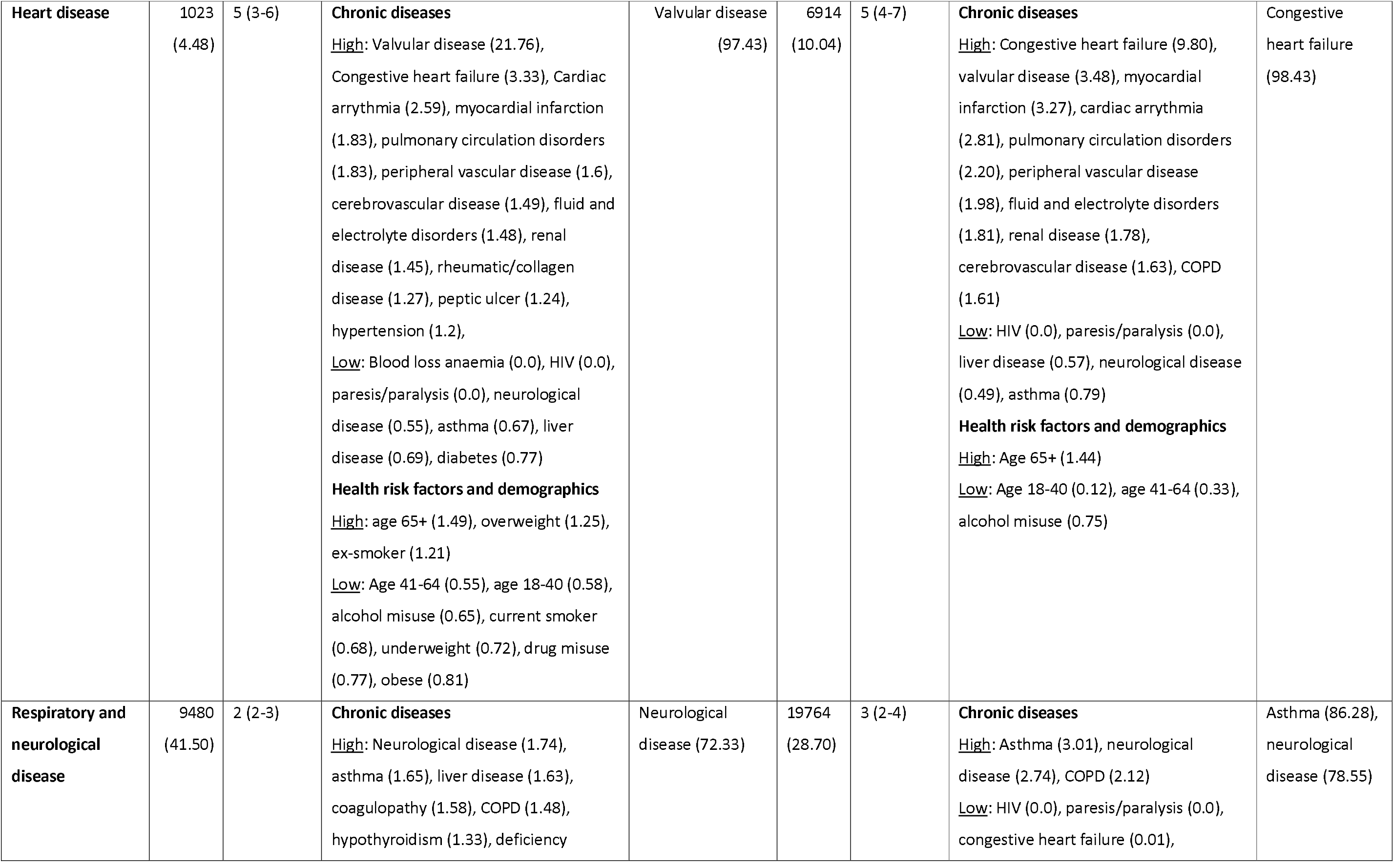
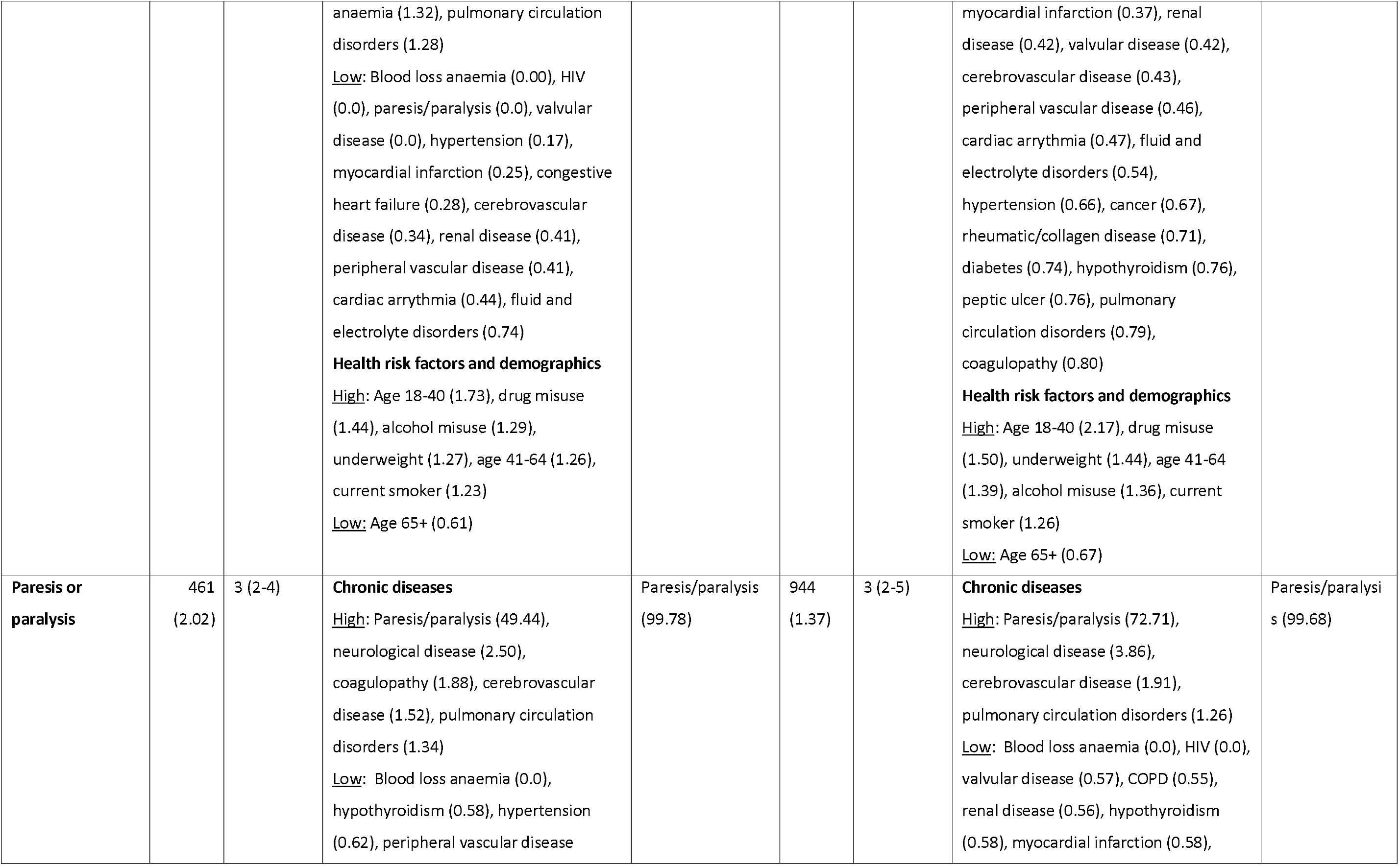
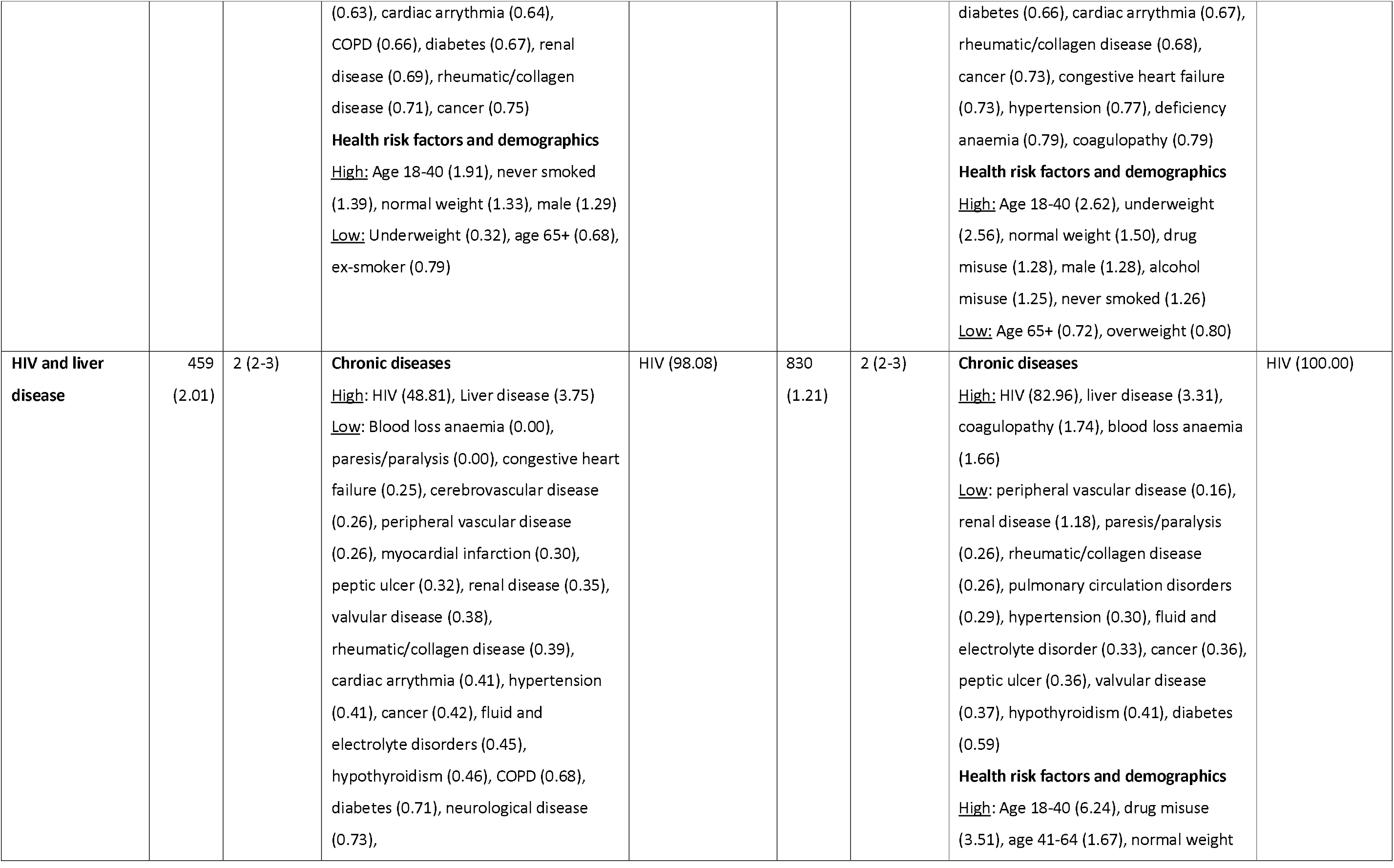
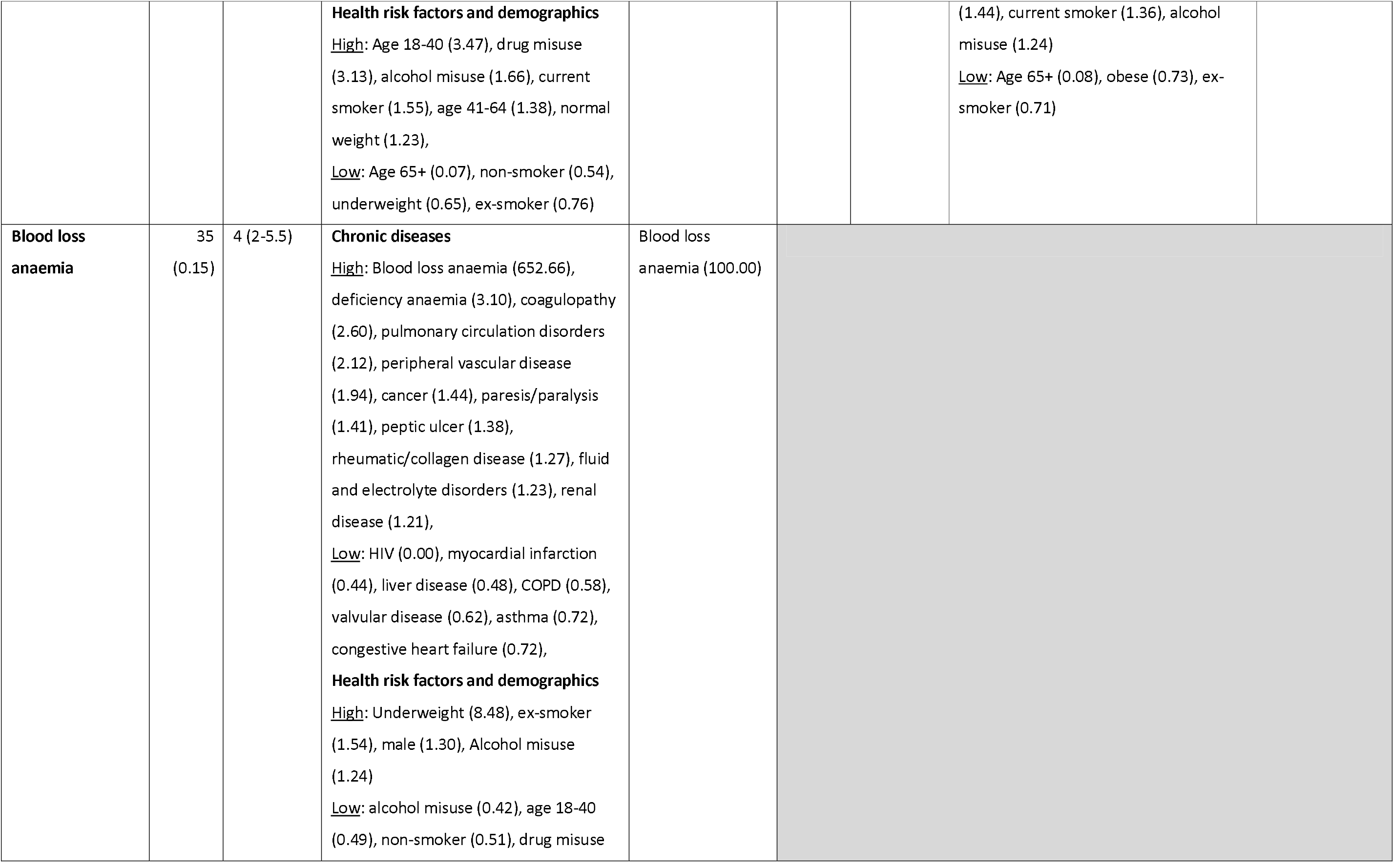
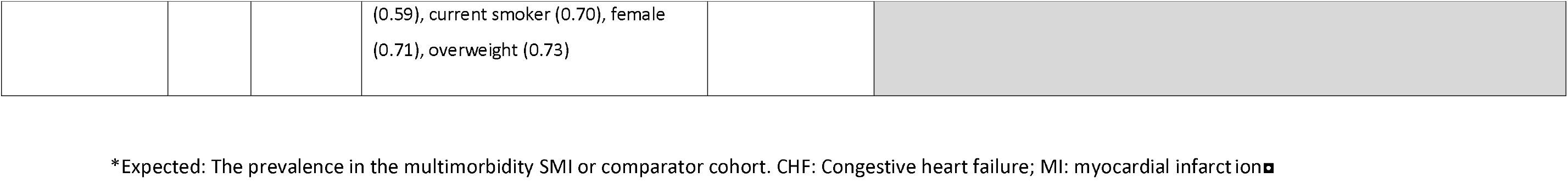
Profiles of multimorbidity identified in K-means cluster analysis.

|  | SMI (n=22,843) |  |  |  | No SMI (n=68,856) |  |  |  |
| --- | --- | --- | --- | --- | --- | --- | --- | --- |
| Cluster description | N (%)<br>in cluster | Median number of physical health conditions (IQR) | Observed in cluster/Expected* > 1.2 or <0.8 | >=70% of those with variable in cluster (%) | N (%)<br>in cluster | Median number of physical health conditions (IQR) | Observed in cluster/Expected* > 1.2 or <0.8 | >=70% of those with variable in cluster (%) |
| Hypertension and varied multimorbidity | 11385 (49.84) | 3 (2-4) | **Chronic diseases**<br><u>High</u>: Hypertension (1.71), myocardial infarction (1.58), cerebrovascular disease (1.52), renal disease (1.49), peripheral vascular disease (1.48), congestive heart failure (1.43), cardiac arrhythmia (1.36),<br><u>Low</u>: Blood loss anaemia (0.0), HIV (0.0), paresis/paralysis (0.0), valvular disease (0.0), neurological disease (0.37), liver disease (0.40), coagulopathy (0.47), asthma (0.48), COPD (0.63), pulmonary circulation disorders (0.68), deficiency anaemia (0.73), hypothyroidism (0.76),<br>**Health risk factors and demographics**<br><u>High</u>: Age 65+ (1.34)<br><u>Low</u>: Age 18-40 (0.3), drug misuse (0.57), alcohol misuse (0.76), age 41-64 (0.81), current smoker (0.82) | Hypertension (85.18), myocardial infarction (78.91), cerebrovascular disease (75.62), renal disease (74.15), peripheral vascular disease (73.57), congestive heart failure (71.26) | 40404 (56.68) | 3 (2-4) | **Chronic diseases**<br><u>High</u>: None<br><u>Low</u>: Congestive heart failure (0.00), paresis/paralysis (0.00), HIV (0.00), asthma (0.05), neurological disease (0.17), COPD (0.37)<br>**Health risk factors and demographics**<br><u>High</u>: None<br><u>Low</u>: Age 18-40 (0.43), underweight (0.74) |  |
| **Heart disease** | 1023<br>(4.48) | 5 (3-6) | **Chronic diseases**<br><u>High</u>: Valvular disease (21.76), Congestive heart failure (3.33), Cardiac arrhythmia (2.59), myocardial infarction (1.83), pulmonary circulation disorders (1.83), peripheral vascular disease (1.6), cerebrovascular disease (1.49), fluid and electrolyte disorders (1.48), renal disease (1.45), rheumatic/collagen disease (1.27), peptic ulcer (1.24), hypertension (1.2),<br><u>Low</u>: Blood loss anaemia (0.0), HIV (0.0), paresis/paralysis (0.0), neurological disease (0.55), asthma (0.67), liver disease (0.69), diabetes (0.77)<br>**Health risk factors and demographics**<br><u>High</u>: age 65+ (1.49), overweight (1.25), ex-smoker (1.21)<br><u>Low</u>: Age 41-64 (0.55), age 18-40 (0.58), alcohol misuse (0.65), current smoker (0.68), underweight (0.72), drug misuse (0.77), obese (0.81) | Valvular disease<br>(97.43) | 6914<br>(10.04) | 5 (4-7) | **Chronic diseases**<br><u>High</u>: Congestive heart failure (9.80), valvular disease (3.48), myocardial infarction (3.27), cardiac arrhythmia (2.81), pulmonary circulation disorders (2.20), peripheral vascular disease (1.98), fluid and electrolyte disorders (1.81), renal disease (1.78), cerebrovascular disease (1.63), COPD (1.61)<br><u>Low</u>: HIV (0.0), paresis/paralysis (0.0), liver disease (0.57), neurological disease (0.49), asthma (0.79)<br>**Health risk factors and demographics**<br><u>High</u>: Age 65+ (1.44)<br><u>Low</u>: Age 18-40 (0.12), age 41-64 (0.33), alcohol misuse (0.75) | Congestive heart failure<br>(98.43) |
| **Respiratory and neurological disease** | 9480<br>(41.50) | 2 (2-3) | **Chronic diseases**<br><u>High</u>: Neurological disease (1.74), asthma (1.65), liver disease (1.63), coagulopathy (1.58), COPD (1.48), hypothyroidism (1.33), deficiency<br>anaemia (1.32), pulmonary circulation disorders (1.28)<br><u>Low:</u> Blood loss anaemia (0.00), HIV (0.0), paresis/paralysis (0.0), valvular disease (0.0), hypertension (0.17), myocardial infarction (0.25), congestive heart failure (0.28), cerebrovascular disease (0.34), renal disease (0.41), peripheral vascular disease (0.41), cardiac arrhythmia (0.44), fluid and electrolyte disorders (0.74)<br>**Health risk factors and demographics**<br><u>High:</u> Age 18-40 (1.73), drug misuse (1.44), alcohol misuse (1.29), underweight (1.27), age 41-64 (1.26), current smoker (1.23)<br><u>Low:</u> Age 65+ (0.61) | Neurological disease (72.33) | 19764<br>(28.70) | 3 (2-4) | **Chronic diseases**<br><u>High</u>: Asthma (3.01), neurological disease (2.74), COPD (2.12)<br><u>Low</u>: HIV (0.0), paresis/paralysis (0.0), congestive heart failure (0.01),<br>myocardial infarction (0.37), renal disease (0.42), valvular disease (0.42), cerebrovascular disease (0.43), peripheral vascular disease (0.46), cardiac arrhythmia (0.47), fluid and electrolyte disorders (0.54), hypertension (0.66), cancer (0.67), rheumatic/collagen disease (0.71), diabetes (0.74), hypothyroidism (0.76), peptic ulcer (0.76), pulmonary circulation disorders (0.79), coagulopathy (0.80)<br>**Health risk factors and demographics**<br><u>High:</u> Age 18-40 (2.17), drug misuse (1.50), underweight (1.44), age 41-64 (1.39), alcohol misuse (1.36), current smoker (1.26)<br><u>Low:</u> Age 65+ (0.67) | Asthma (86.28), neurological disease (78.55) |
| **Paresis or paralysis** | 461 (2.02) | 3 (2-4) | **Chronic diseases**<br><u>High:</u> Paresis/paralysis (49.44), neurological disease (2.50), coagulopathy (1.88), cerebrovascular disease (1.52), pulmonary circulation disorders (1.34)<br><u>Low:</u> Blood loss anaemia (0.0), hypothyroidism (0.58), hypertension (0.62), peripheral vascular disease | Paresis/paralysis (99.78) | 944 (1.37) | 3 (2-5) | **Chronic diseases**<br><u>High:</u> Paresis/paralysis (72.71), neurological disease (3.86), cerebrovascular disease (1.91), pulmonary circulation disorders (1.26)<br><u>Low:</u> Blood loss anaemia (0.0), HIV (0.0), valvular disease (0.57), COPD (0.55), renal disease (0.56), hypothyroidism (0.58), myocardial infarction (0.58), | Paresis/paralysis (99.68) |
|  |  |  | (0.63), cardiac arrhythmia (0.64), COPD (0.66), diabetes (0.67), renal disease (0.69), rheumatic/collagen disease (0.71), cancer (0.75)<br>**Health risk factors and demographics**<br><u>High</u>: Age 18-40 (1.91), never smoked (1.39), normal weight (1.33), male (1.29)<br><u>Low</u>: Underweight (0.32), age 65+ (0.68), ex-smoker (0.79) |  |  |  | diabetes (0.66), cardiac arrhythmia (0.67), rheumatic/collagen disease (0.68), cancer (0.73), congestive heart failure (0.73), hypertension (0.77), deficiency anaemia (0.79), coagulopathy (0.79)<br>**Health risk factors and demographics**<br><u>High</u>: Age 18-40 (2.62), underweight (2.56), normal weight (1.50), drug misuse (1.28), male (1.28), alcohol misuse (1.25), never smoked (1.26)<br><u>Low</u>: Age 65+ (0.72), overweight (0.80) |  |
| **HIV and liver disease** | 459<br>(2.01) | 2 (2-3) | **Chronic diseases**<br><u>High</u>: HIV (48.81), Liver disease (3.75)<br><u>Low</u>: Blood loss anaemia (0.00), paresis/paralysis (0.00), congestive heart failure (0.25), cerebrovascular disease (0.26), peripheral vascular disease (0.26), myocardial infarction (0.30), peptic ulcer (0.32), renal disease (0.35), valvular disease (0.38), rheumatic/collagen disease (0.39), cardiac arrhythmia (0.41), hypertension (0.41), cancer (0.42), fluid and electrolyte disorders (0.45), hypothyroidism (0.46), COPD (0.68), diabetes (0.71), neurological disease (0.73), | HIV (98.08) | 830<br>(1.21) | 2 (2-3) | **Chronic diseases**<br><u>High</u>: HIV (82.96), liver disease (3.31), coagulopathy (1.74), blood loss anaemia (1.66)<br><u>Low</u>: peripheral vascular disease (0.16), renal disease (1.18), paresis/paralysis (0.26), rheumatic/collagen disease (0.26), pulmonary circulation disorders (0.29), hypertension (0.30), fluid and electrolyte disorder (0.33), cancer (0.36), peptic ulcer (0.36), valvular disease (0.37), hypothyroidism (0.41), diabetes (0.59)<br>**Health risk factors and demographics**<br><u>High</u>: Age 18-40 (6.24), drug misuse (3.51), age 41-64 (1.67), normal weight | HIV (100.00) |
|  |  |  | **Health risk factors and demographics**<br><u>High</u>: Age 18-40 (3.47), drug misuse (3.13), alcohol misuse (1.66), current smoker (1.55), age 41-64 (1.38), normal weight (1.23),<br><u>Low</u>: Age 65+ (0.07), non-smoker (0.54), underweight (0.65), ex-smoker (0.76) |  |  |  | (1.44), current smoker (1.36), alcohol misuse (1.24)<br><u>Low</u>: Age 65+ (0.08), obese (0.73), ex-smoker (0.71) |  |
| **Blood loss anaemia** | 35<br>(0.15) | 4 (2-5.5) | **Chronic diseases**<br><u>High</u>: Blood loss anaemia (652.66), deficiency anaemia (3.10), coagulopathy (2.60), pulmonary circulation disorders (2.12), peripheral vascular disease (1.94), cancer (1.44), paresis/paralysis (1.41), peptic ulcer (1.38), rheumatic/collagen disease (1.27), fluid and electrolyte disorders (1.23), renal disease (1.21),<br><u>Low</u>: HIV (0.00), myocardial infarction (0.44), liver disease (0.48), COPD (0.58), valvular disease (0.62), asthma (0.72), congestive heart failure (0.72),<br>**Health risk factors and demographics**<br><u>High</u>: Underweight (8.48), ex-smoker (1.54), male (1.30), Alcohol misuse (1.24)<br><u>Low</u>: alcohol misuse (0.42), age 18-40 (0.49), non-smoker (0.51), drug misuse | Blood loss anaemia (100.00) |  |  |  |  |

|  |  |  |  |
| --- | --- | --- | --- |
|  |  |  | (0.59), current smoker (0.70), female<br>(0.71), overweight (0.73) |

\*Expected: The prevalence in the multimorbidity SMI or comparator cohort. CHF: Congestive heart failure; MI: myocardial infarction■

**Figure 4:**
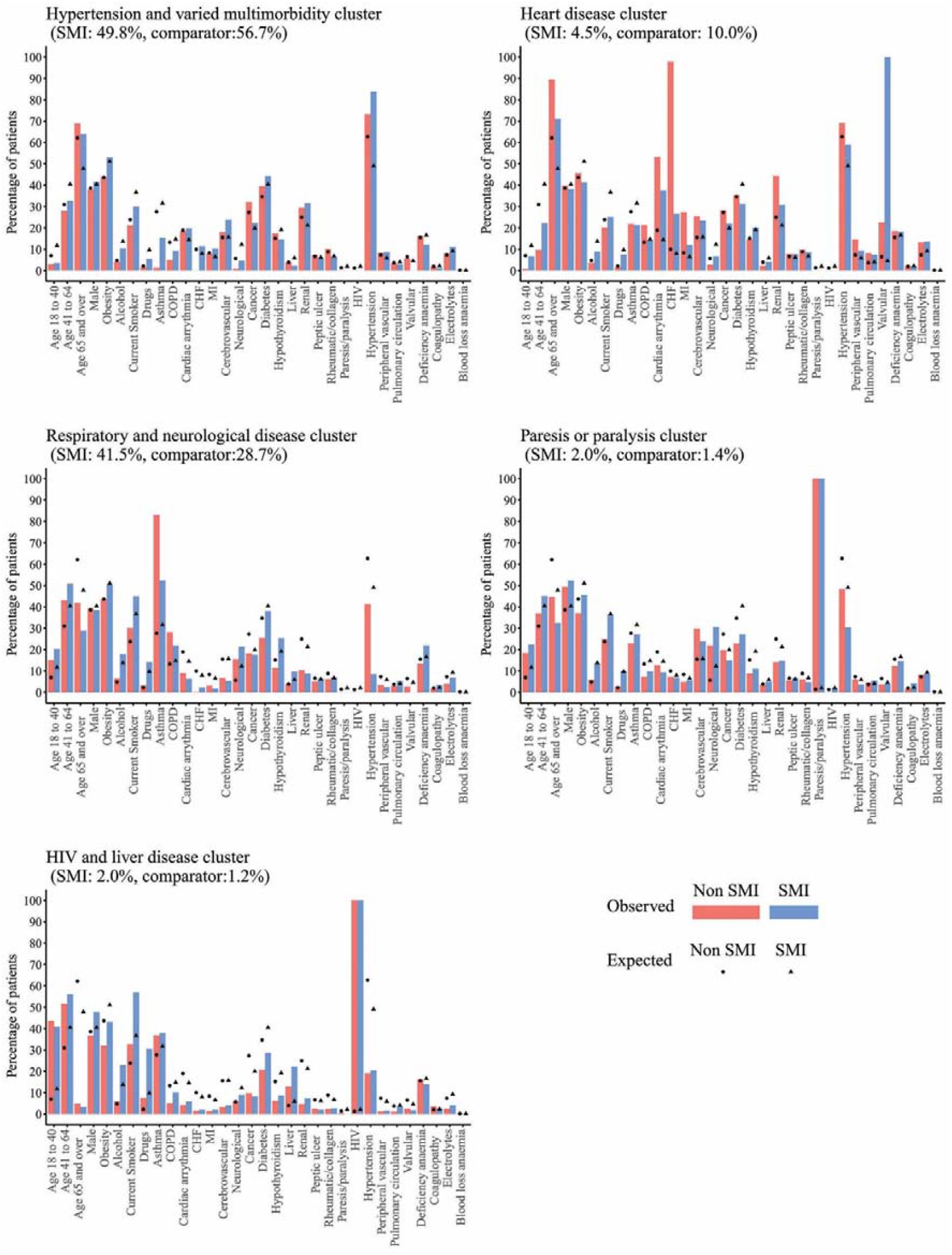
Prevalence of physical health conditions, health risk factors and demographics in the SMI and comparator cohorts for the three most common multimorbidity clusters observed in the SMI cohort.

Patients with SMI were more likely to be classified in the “respiratory and neurological” (SMI: 41.5%, comparator: 28.7%) or “HIV and liver disease” clusters. These clusters had a higher prevalence of health risk factors and more patients aged under 65 than expected (Table 3). Patients with SMI were also more likely to fall in the “paresis or paralysis” cluster, also defined by younger age, but without an increased prevalence of health risk factors, and with an increased prevalence of male patients.

When we included health risk factors in the cluster analysis, the largest cluster we identified was a “general multimorbidity” cluster; accounting for 63.4% of the SMI and 53.0% of the comparator cohorts. We also identified a cluster defined by a high prevalence of health risk factors, male gender, liver disease, HIV and younger age. This cluster accounted for 16.4% of patients with SMI and 2.2% of patients in the comparator cohort.

### Sensitivity analyses

In sensitivity analysis, recoding missing ethnicity to “missing” did not alter the interpretation of disease prevalence (Supplementary table 1).

## Discussion

Our study investigated physical health conditions and multimorbidity in a large cohort of patients with SMI and matched comparators. Clustering of multimorbid health conditions was not dramatically different between those with and without SMI, despite higher prevalence of many physical health conditions in the SMI cohort. However, patients with a diagnosis of SMI developed multimorbidity earlier and were more likely to fall into a cluster of “respiratory and neurological disease” cluster, than comparators.

### Patterns of physical health conditions and multimorbidity

In line with other studies (16, 18, 47, 48), we found an increased prevalence of multimorbidity in women in both SMI and comparator cohorts. While increased prevalence of multimorbidity with increasing age is well described in the general population (9, 33, 48), we found the largest differences between those with and without SMI in the young and middle age groups, suggesting those with SMI develop multimorbidity earlier than the general population. The high premature mortality rate in patients with SMI(49), in part driven by poor physical health, may reduce the prevalence of older-age multimorbidity in this population.

Similarities in physical health profiles of those with and without SMI were apparent in individual and disease-pair ranking, MCA and cluster analysis. Two previous studies have found similarity in ranking the most frequently diagnosed conditions and pairs of conditions between those with and without SMI (1, 15), and a hospital-based study of self-reported physical health conditions in 1060 psychiatric patients and 837 members of the general population, found similar profiles of multimorbidity between the two cohorts using Latent Class Analysis (19). Our analysis suggests that, despite these similarities, there is a large group of patients with SMI developing multimorbidity at a younger age than the general population, and with respiratory or neurological diseases rather than cardiovascular diseases.

Health risk factors likely explain some of the increased risk of physical health conditions and multimorbidity in people with SMI. We found that including smoking status, BMI category, and alcohol and drug misuse in cluster analysis resulted in a higher proportion of patients in the SMI cohort being in a “health risk” cluster, and that adjusting for these factors decreased the odds ratios of physical health conditions between SMI and comparator cohorts, particularly for liver disease, HIV, COPD, diabetes and hypertension. A previous study found that substance misuse was associated with higher prevalence of hepatitis and HIV(50), while the links between COPD and smoking and diabetes, hypertension and obesity are well established. Interventions to modify these risk factors, for example via smoking or alcohol cessation support(51, 52), have been shown to be effective in people with SMI and need to be more widely available.

### Under-ascertainment of physical health conditions in patients with schizophrenia

Our study found that patients with SMI had increased odds of 19 of the 24 studied conditions compared to comparators. However, when stratified by diagnosis, we identified lower prevalence of a range of physical health conditions in patients with schizophrenia. This is surprising given the observed high prevalence of smoking, obesity, and alcohol and drug misuse in this group, and known side effects of antipsychotic medication(10). Under-reporting is also likely not due to lack of contact between primary care physicians and patients with SMI, as annual health checks in primary care have been recommended and incentivised in this patient group since 2004. Lower prevalence of cardiovascular disease(16, 18, 53, 54) and cancer(18) have been reported in other studies using routine primary care data, and our study corroborates this finding using a matched comparator population and controlling for both demographic and health risk factors. This could reflect poor access to care, under-diagnosis, or diagnostic overshadowing in the schizophrenia population. There is evidence that those with schizophrenia are more likely to have physical health conditions recorded at the time of death (39, 55), suggesting late and missed diagnoses in this population.

### Strengths and limitations

To our knowledge this study is the largest investigation of multimorbidity, and clustering of physical health conditions in patients with SMI. A key strength of our study was the ability to adjust for smoking, BMI category, alcohol, and drug misuse as risk factors for physical health conditions.

The large sample size of this study, and representativeness of data from CPRD(34, 35), suggests the results of this study are generalisable to the UK population. The prevalence of multimorbidity in the SMI cohort was similar to previous studies(16, 18, 56), however prevalence of multimorbidity depends on the conditions studied and therefore other studies have found substantially different(15, 17) estimates. As with all studies using electronic health records, a limitation of this study is potential biases in recording variables. While the apparent under-recording of a range of physical health conditions in those with schizophrenia is a clinically important finding, it limits the interpretation of disease prevalence and multimorbidity clusters in this population.

There may be residual confounding due to missing information, however missing values for smoking status, ethnicity and BMI were replaced in line with other primary care studies(42, 43), and sensitivity analyses performed for ethnicity. Alcohol misuse was based on medical code lists and did not account for the level of alcohol consumption, nor include patients that had consumption recorded without an accompanying alcohol misuse code. While we were unable to control for deprivation, patients were matched on GP practice and therefore from a broadly comparable geographic area.

This study focused on physical health conditions ever diagnosed, which limits the study of temporality of diagnoses of SMI and physical health conditions. However, with both SMI and chronic physical health conditions, a prodromal stage or period of undiagnosed disease may occur and therefore diagnosis dates may not give a clear indication of temporal association. Furthermore, previous studies have found increased physical health problems prior to schizophrenia(57, 58) and bipolar disorder diagnoses(59) and that health risk factors for physical health conditions such as smoking(60) and alcohol and drug misuse may be present prior to SMI diagnosis(61).

## Conclusions

We found that there is an increased prevalence of physical health conditions and physical health multimorbidity in those with SMI, but that these conditions cluster in people with SMI in a similar manner to the general population. The absence of novel clusters of disease suggests that the same drivers of physical health conditions are at play in both populations, and therefore research and service provision for patients with SMI should focus on the same risk factors and disease clusters as in the general population. However, while much of the focus of multimorbidity in the general population has been on old age, our study found that multimorbidity starts earlier in those with SMI. Crucially, our study found that those with SMI were more likely to fall into clusters of disease characterised by asthma, COPD and neurological diseases such as epilepsy or multiple sclerosis, than the comparator cohort, and that health risk factors such as smoking, drug or alcohol misuse play a larger role. This highlights highlighting an unmet need in terms of interventions aimed at a younger cohort of multimorbid patients and demonstrating the importance of physical health checks in this population.

Further work is warranted to investigate the temporality of SMI and physical health condition diagnoses, and trajectories of multimorbidity in this population. The low prevalence of some physical health conditions in the schizophrenia cohort also requires further investigation, to elucidate the reasons for this finding. Finally, the relevance of the identified clusters to outcomes such as hospitalisation and mortality, both in patients with and without SMI, is an area for future research.

## Data Availability

No additional data are available.

## Funding

This study was supported by Public Health England (PhD2019/002), the Wellcome Trust (211085/Z/18/Z), the Medical Research Council (MC\PC\17216), University College London Hospitals NIHR Biomedical Research Centre and the NIHR ARC North Thames Academy. The funders had no role in study design; or conduct; in the writing of the report; or in the decision to submit the article for publication

## Notes

### Competing Interest Statement

The authors have declared no competing interest.

### Author Declarations

Ethical approval for this study was obtained from the Independent Scientific Advisory Committee of CPRD (protocol no. 18_288).

