## Supplementary data for "Clustering of physical health multimorbidity in 68,392 people with severe mental illness and matched comparators: a lifetime prevalence analysis of United Kingdom primary care data"

#### Supplementary figure 1: Pattern of variables in Multiple Correspondence Analysis in SMI and comparator cohorts

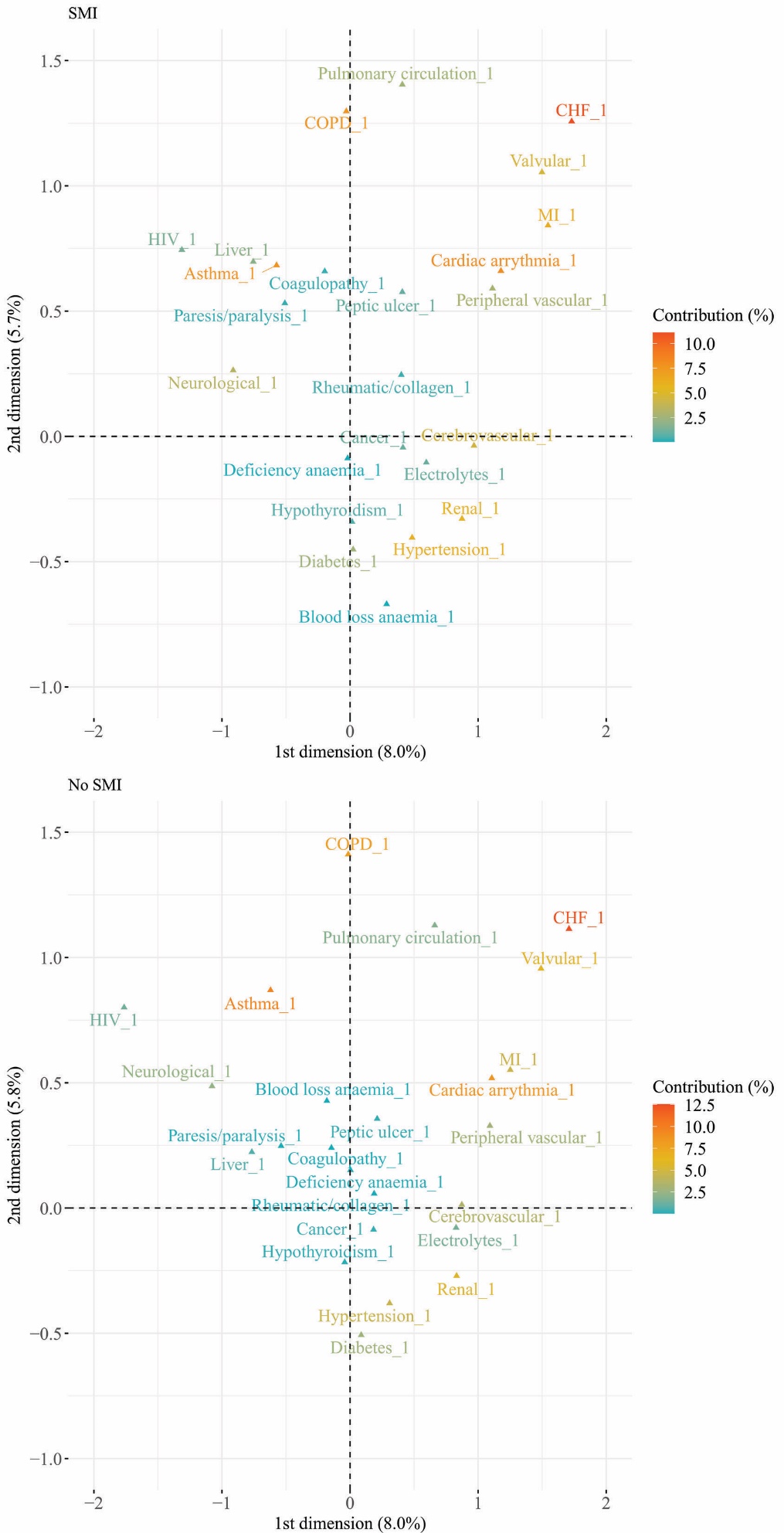

#### Supplementary table 1: Fully adjusted odds ratios of physical health conditions in patients with missing ethnicity, compared to other ethnicities

| **Condition** | **OR (95%CI)** |
| --- | --- |
| Asthma | 0.83(0.81-0.84) |
| COPD | 0.72(0.69-0.75) |
| Cardiac arrythmia | 0.80(0.78-0.83) |
| Congestive heart failure | 1.02(0.97-1.07) |
| Myocardial infarction | 1.00(0.95-1.06) |
| Cerebrovascular disease | 0.97(0.94-1.01) |
| Neurological disease | 0.84(0.81-0.88) |
| Cancer | 0.87(0.85-0.90) |
| Diabetes | 0.74(0.72-0.76) |
| Hypothyroidism | 0.80(0.77-0.83) |
| Liver disease | 0.89(0.84-0.94) |
| Renal disease | 0.67(0.65-0.69) |
| Peptic ulcer | 0.96(0.91-1.01) |
| Rheumatic/collagen diseases | 0.90(0.85-0.94) |
| Paresis/paralysis | 0.90(0.81-0.99) |
| HIV | 0.63(0.57-0.69) |
| Hypertension | 0.81(0.79-0.83) |
| Peripheral vascular disease | 0.98(0.93-1.04) |
| Pulmonary circulation disorders | 0.88(0.82-0.94) |
| Valvular disease | 0.75(0.71-0.80) |
| Deficiency anaemia | 0.82(0.79-0.85) |
| Blood loss anaemia | 0.94(0.69-1.29) |
| Coagulopathy | 0.85(0.77-0.93) |
| Fluid and electrolyte disorders | 0.74(0.70-0.78) |

#### Supplementary table 2: Prevalence of multimorbidity by age at end of follow up

|  | **Prevalence, n, (%)** | | | | | **Partially adjusted, Odds Ratio (95% CI)** | | | | **Fully adjusted, Odds Ratio (95% CI)** | | | |
| --- | --- | --- | --- | --- | --- | --- | --- | --- | --- | --- | --- | --- | --- |
|  | **No SMI** | **SMI** | **Schiz** | **Bipolar** | **Other** | **SMI** | **Schiz** | **Bipolar** | **Other** | **SMI** | **Schiz** | **Bipolar** | **Other** |
| Age at end of follow up |  |  |  |  |  |  |  |  |  |  |  |  |  |
| 20-29 | 1318 (3.1) | 722 (7.2) | 143 (6.5) | 230 (7.7) | 349 (7.3) | 2.5 (2.3-2.7) | 2.4 (2.0-2.8) | 2.4 (2.1-2.8) | 2.6 (2.3-2.9) | 2.0 (1.8-2.2) | 1.8 (1.5-2.2) | 1.9 (1.6-2.2) | 2.1 (1.9-2.4) |
| 30-39 | 3074 (5.5) | 1708 (12.3) | 396 (11.3) | 605 (12.6) | 707 (12.6) | 2.4 (2.3-2.6) | 2.4 (2.2-2.7) | 2.2 (2.-2.4) | 2.6 (2.4-2.8) | 1.6 (1.5-1.7) | 1.6 (1.4-1.7) | 1.5 (1.3-1.6) | 1.8 (1.6-2.) |
| 40-49 | 5701 (11.3) | 3047 (22.9) | 689 (21.3) | 1237 (24.5) | 1121 (22.4) | 2.3 (2.2-2.4) | 2.2 (2.0-2.4) | 2.5 (2.3-2.6) | 2.3 (2.1-2.5) | 1.5 (1.5-1.6) | 1.4 (1.3-1.6) | 1.6 (1.5-1.7) | 1.5 (1.4-1.7) |
| 50-59 | 10030 (22.9) | 4331 (38.2) | 921 (37.7) | 1808 (38.7) | 1602 (38.0) | 2.1 (2.0-2.2) | 2.0 (1.8-2.2) | 2.1 (2.-2.3) | 2.0 (1.9-2.2) | 1.5 (1.4-1.6) | 1.4 (1.3-1.6) | 1.5 (1.4-1.6) | 1.5 (1.4-1.6) |
| 60-69 | 12133 (41.4) | 4030 (54.2) | 762 (48.3) | 1725 (56.9) | 1543 (54.7) | 1.7 (1.6-1.8) | 1.3 (1.2-1.4) | 1.9 (1.8-2.1) | 1.7 (1.5-1.8) | 1.3 (1.3-1.4) | 1.0 (0.9-1.1) | 1.5 (1.3-1.6) | 1.4 (1.3-1.5) |
| 70-79 | 14031 (63.8) | 3866 (68.9) | 655 (62.1) | 1585 (73.1) | 1626 (68.3) | 1.3 (1.2-1.3) | 0.9 (0.8-1.0) | 1.6 (1.4-1.7) | 1.2 (1.1-1.3) | 1.2 (1.1-1.3) | 0.9 (0.8-1.0) | 1.4 (1.3-1.6) | 1.2 (1.1-1.3) |
| 80-89 | 15464 (78.3) | 3709 (76.5) | 483 (67.8) | 1072 (80.5) | 2154 (76.8) | 0.9 (0.8-1.0) | 0.6 (0.5-0.7) | 1.2 (1.0-1.3) | 0.9 (0.8-1.0) | 0.9 (0.9-1.0) | 0.6 (0.5-0.7) | 1.1 (1.-1.3) | 1.0 (0.9-1.1) |
| 90-99 | 6775 (79.2) | 1356 (76.1) | 130 (69.1) | 186 (75.3) | 1040 (77.2) | 0.8 (0.7-0.9) | 0.6 (0.4-0.8) | 0.8 (0.6-1.1) | 0.9 (0.8-1.0) | 0.9 (0.8-1.0) | 0.5 (0.4-0.7) | 0.8 (0.6-1.1) | 0.9 (0.8-1.1) |
